# ‘Small Steps’ towards improving 24-hour time-use behaviours to decrease the risk of dementia: protocol for a personalised, web-based randomised controlled trial in community-dwelling older adults

**DOI:** 10.1101/2025.05.26.25328336

**Authors:** Maddison L Mellow, Henry T Blake, Ty Ferguson, Bethany Robins, Dorothea Dumuid, Timothy Olds, Ty Stanford, Kate Laver, Hannah AD Keage, Alison M Coates, Alexandra T Wade, Michelle Rogers, Aaron Davis, Lui Di Venuto, Emma Tregoweth, Catherine Yandell, Barbara Tainsh, Ashleigh E Smith

## Abstract

**Introduction:** Addressing physical inactivity is a promising dementia risk reduction strategy due to its direct benefits for brain health, and indirect benefits for other modifiable dementia risk factors. A potential limitation of previous interventions is that they often overlook how increasing physical activity affects other behaviours throughout the 24-hour day, such as sleep and sedentary behaviour, which are also important for brain health. Further, interventions are rarely tailored to the individual, considering their needs, preferences and constraints that may serve as barriers or facilitators to behaviour change. The current phase I randomised controlled trial, *Small Steps*, aims to investigate feasibility, acceptability and preliminary effectiveness of a personalised 24-hr time-use intervention to improve lifestyle and cognitive health in older adults.

**Methods and analysis:** Participants aged ≥65 years from Adelaide, South Australia will be recruited and randomised to either the Extended or Condensed program. During the first 12 weeks, participants in the Extended program will use a tailored website to set personalised weekly goals to move towards their ‘optimal’ 24-hr day for brain health, facilitated by weekly website ‘check-ins’ and weekly phone calls with a research staff member. Participants randomised to the Condensed program will have access to the website educational resources only but will not undergo personalised goal setting or telephone calls. Following the introductory phase (first 12 weeks), phone calls will be gradually withdrawn for the Extended program. Primary (feasibility and acceptability) and secondary outcomes (changes in time use, cognitive function and behaviour change metrics) will be assessed 12, 24 and 36 weeks after the beginning of the intervention.

**Conclusions:** Given the major and growing financial and social burdens associated with dementia, if successful, this trial will be the first to facilitate effective knowledge translation and personalisation of 24-hr time-use intervention approaches for dementia risk-reduction.

**Ethics and dissemination:** Ethics approval has been obtained from the University of South Australia’s Human Research Ethics Committee (205989). Study findings will be disseminated through peer-reviewed journal articles, conference presentations, media releases and community engagement.

**Trial registration number:** The clinical trial protocol was registered at ClinicalTrials.gov (unique protocol ID: NCT06291909).

**STRENGTHS AND LIMITATIONS OF THIS STUDY:**

- The *Small Steps* study will investigate the feasibility, acceptability and preliminary effectiveness of a personalised 24-hour time-use intervention to improve lifestyle and cognitive health in 88 community-dwelling older adults.
- Participants randomised to the Extended program (intervention) will be supported to improve their physical activity, sleep and sedentary behaviour patterns for 12 weeks with weekly researcher phone calls and website check-ins.
- Intervention goals will be personalised to the individual based on their current time use, projected ‘optimal’ time use, and their preferences and constraints associated with 24-hr time use.
- The intervention was co-designed with a separate cohort of community-dwelling older adults and allied health professionals.
- Recruitment will be based on self-report physical activity levels, which may result in recruitment of participants who have underestimated their activity levels and limit the generalisability of the findings.

## INTRODUCTION

Engaging in sufficient physical activity (PA) is a promising dementia prevention strategy due to its direct impact on brain health, and indirect benefits by improving other risk factors including obesity, diabetes, social isolation, and depression (1). Achieving sufficient levels of PA is associated with larger brain volumes, greater cortical thickness, increased cortical connectivity between functionally relevant brain regions and facilitation of synaptic neuroplasticity (2, 3). Whilst multi-domain dementia prevention trials that target physical activity in combination with other lifestyle approaches (e.g., targeting cardiovascular risk factors or poor diet (4)) have shown promise, achieving longer term changes in behaviour remains a key challenge for PA interventions in older adult populations (5). Designing sustainable interventions which promote the maintenance of new increases in PA beyond the end of the intervention period is therefore a key challenge for dementia prevention moving forwards.

### Taking a “24-hour” approach to dementia prevention

One potential limitation of previous interventions which have targeted PA is that they have not considered the impact of increasing time in PA on the rest of the 24-hour day. PA does not exist in isolation but is one of three “time-use behaviours”, alongside sleep and sedentary behaviour (i.e., sitting) (6). These three time-use behaviours are mutually exclusive and exhaustive components of the 24-hour day (6). Thus, increasing time in PA as part of an intervention must result in a decrease in one or both of the remaining behaviours. This reallocation of time likely has important implications for brain health outcomes, as all three behaviours are associated with brain health in older adulthood (2, 7).

Observational studies have suggested an important role for PA, particularly moderate-to-vigorous PA (MVPA), in promoting brain health and function in older adults (8, 9). A previous review on the associations between combinations of PA, sleep and sedentary behaviour and cognitive function in older adults identified that increasing time in MVPA at the expense of sedentary behaviour results in the greatest benefits (10). However, not all sedentary behaviours are equal in their relationship with cognitive function (e.g., TV watching is detrimental for cognitive function, whilst reading and computer use is beneficial (11, 12)).

Sleep duration is widely thought to share an inverted U-shaped relationship with brain health and function, where long or short sleep durations (e.g., less than 6 or more than 9 hours) are associated with poorer outcomes, although findings are mixed (13, 14). Moreover, sleep (duration and quality) may moderate PA or exercise-induced cognitive change (15). Taken together, it is likely important to optimise the *balance* of PA, sleep and sedentary behaviour across the 24-hour day, or *24-hr time-use composition*, for brain health rather than targeting these behaviours in isolation. Whilst guidelines for 24-hour movement behaviours to promote overall health do exist (16), to our knowledge there are no intervention studies which have targeted the 24-hour day holistically for dementia prevention.

### Personalising 24-hour time-use interventions for brain health

Whilst there has been a rapid increase in observational research studying relationships between 24-hour time use and brain health in the past 5 years, the impact of personal characteristics (e.g., health and sociodemographic factors) and social determinants of health on these associations has been largely overlooked (8, 17). In the context of a 24-hr time-use intervention for dementia prevention, a participant’s unique combination of non-modifiable (e.g., age, sex, genetics) and modifiable dementia risk factors (e.g., cardiovascular health, obesity, smoking), as well as their current levels of PA, sleep and sedentary behaviour, may alter the ‘optimal’ 24-hr time-use composition, and thus the intervention strategy, needed to improve brain health and reduce dementia risk (17, 18). Moreover, ‘one-size-fits-all’ interventions which do not consider the unique time constraints and preferences of the individual may create barriers to intervention adherence and success.

Taking a personalised approach to optimising 24-hour time use may result in greater brain health benefits and facilitate the maintenance of behaviour change beyond the end of the intervention period. However, delivering such an intervention presents a number of potential challenges. First, personalised intervention strategies may be highly varied between participants due to their unique needs, preferences and constraints. Second, a tailored 24-hr time-use intervention may involve complex behaviour change patterns which vary week-to-week (e.g., exchanging sleep for PA one week, and sitting for PA in another week). Digital technology interventions may help to overcome such challenges, as well as barriers to adherence such as time constraints or geographical location of participants. These interventions have demonstrated considerable efficacy for a number of health behaviours including substance use, diet and physical activity (19). Moreover, using custom websites to translate potentially complex interventions (e.g., reallocating time between behaviours within the 24-hr day) to accessible and relatable instructions may also promote autonomy, motivation and enjoyment. However, to ensure appropriateness of delivered intervention level information, complex interventions should be co-designed with the population for which they will be tested on. The current study, *Small Steps*, was co-designed with older adults and health care professionals with results of the co-design process and design decisions detailed elsewhere (20).

## STUDY AIMS

*Small Steps* aims to examine the feasibility, acceptability and preliminary effectiveness of a personalised behavioural intervention to optimise daily PA, sedentary behaviour and sleep patterns (24-hour time-use composition) for cognitive health in older adults. We hypothesise *Small Steps* will be feasible and acceptable, and result in increased time spent in PA (measured from enrolment to the end of the introductory phase) and improved cognitive functioning, compared to generic health advice.

## METHODS

### Ethics and registration

Ethics approval was obtained from the University of South Australia Human Research Ethics Committee (205989). All procedures will be conducted in accordance with the Declaration of Helsinki. The clinical trial protocol was registered at ClinicalTrials.gov (unique protocol ID: NCT06291909) in February 2024, and will be updated following ethical approval of protocol modifications as needed (current version:4, updated in December 2024).

### Study design

*Small Steps* is a two-arm phase I randomised controlled trial (RCT) comparing the feasibility, acceptability and preliminary effectiveness of the Extended program (intervention group) and the Condensed program (control group) on 24-hour time-use patterns in community-dwelling older adults.

### Participants and recruitment

Eighty-eight community-dwelling older adults will be targeted for recruitment from Adelaide and surrounds (South Australia). An *a priori* total sample size of n=88 (44 per group) was calculated to achieve greater than 95% power at the α=0.05 level of significance to detect an additionally beneficial increase (should it exist) of 20 minutes of PA per day on average in the Extended program, compared to the Condensed program, assuming a worst-case attrition of 20%. Empirical power calculations *via* simulation, accounting for bounded, non-negative response values (24-hr time use), were additionally verified using analytical multiple linear regression power analysis methods available in G*Power 3.1.9.7.

Participants will be aged 65 years or older, ambulatory, fluent in the English language and will have access to, and competency in, using digital technology (i.e., mobile phone, tablet, computer). Participants will be screened for the following inclusion criteria: not currently meeting the Australian Physical Activity Guidelines (21) of 150 minutes per week of moderate intensity exercise; deemed safe to participate in PA as per the Exercise Sport Science Australia Adult Pre-Exercise Screening System (22), or through clearance from a health professional; do not score below the mild cognitive impairment (MCI) cut-off on the Telephone Montreal Cognitive Assessment (T-MoCA; 13/22) (23); do not have a current diagnosis of dementia; do not have a known major physical or intellectual disability; do not have a major neurological or psychiatric diagnosis; are not involved in another intervention involving changes to PA or diet, or brain training; do not have vision problems which may prevent them from reading a computer and/or phone screen; are willing and able to provide written informed consent. Within 3 months of study commencement, participants who report a change in medication that might influence outcomes will also be excluded.

Participants will be recruited through *Small Steps* partners (e.g., City of Onkaparinga Council and ACH Group) as well as social media, radio, print media, and flyers distributed by community partner organisations in the greater Adelaide region. Initial eligibility screening will be completed online using a survey in REDCap (24). Interested participants will be asked to provide basic information including name, age, sex, whether they are a resident of the targeted Adelaide suburban areas, and will be screened for eligibility criteria listed above. Finally, a member of the research team will contact interested participants to undertake the T-MoCA.

### Patient and public involvement

Prior to the application for funding, community consultation interviews were conducted with 19 older adults (14 female) and 5 health professionals (Sept – Dec 2021) to inform the design of *Small Steps*. Most older adults revealed they would use a technology delivered *Small Steps* intervention (number of older adults responding positively = 16), providing feedback such as *“…I think, it would be a really well accessed app”* (older adult). Other participants highlighted the need for primary care and allied health dissemination: *“…I think the greatest driver would be your GP”* (older adult). Health professionals shared similar positive comments about the potential of a digital tool describing the need for support and accountability for users: *“…you need that either one-on-one or group support where they can check in, and be slightly accountable, or have that personal feedback”.* Following funding acquisition, *Small Steps* was fully co-designed with older adults and allied health professionals (see elsewhere for detailed description of *Small Steps* co-design process (20).

### Participant randomisation and blinding protocol

Participants will be randomly allocated to treatment via imbalance minimisation using previously identified participant prognostic factors (categorized): age (<75 or ≥75 years), sex (male or female), PA level (average daily steps from Fitbit watch; <3000, 3000-5999, or ≥6000), and self-efficacy for PA score (<51 or ≥51). In the case where participants have shared living arrangements (e.g., if 2 participants live in same household), subsequent participants from the same household will be allocated to the same group as the first randomised participant. Imbalance minimisation was chosen as it has the advantage of achieving a balance of outcome measures between groups at baseline in studies with small sample sizes, which is particularly important where there may be strong prognostic factors and modest treatment effects. Participants will be allocated at a 1:1 ratio of treatment (Extended program) to control (Condensed program). Randomisation will be completed by a separate investigator (TS) who is not involved in data collection. To ensure statistical blinding, generic group allocation labels will be generated by the trial statistician (TS; ‘Group A’ and ‘Group B’), and only staff involved in the intervention delivery (BR, MM, and AS) will know generic label to intervention delivery translation. Participants will be instructed not to discuss their allocation with the researchers during data collection visits.

### Primary outcome measures

Regardless of group assignment (Extended or Condensed program), participants will attend in-person baseline, 12-week, 24-week and 36-week assessment visits where primary and secondary outcome measures will be collected (See Figure 1). Table 1 indicates the data collection schedule for primary and secondary outcomes.

**Figure 1.**
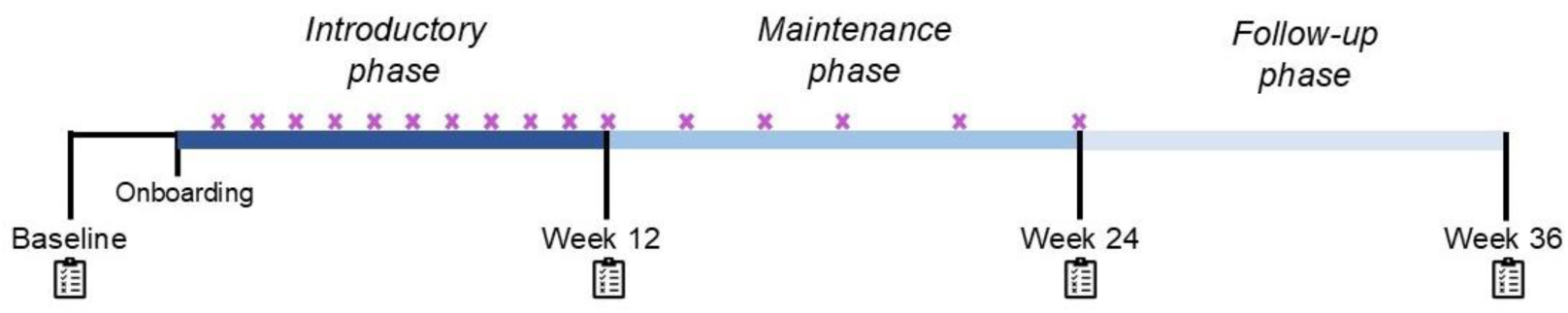
Schematic diagram of study visit schedule for *Small Steps*. Crosses indicate research team check-in phone calls. Phone calls are undertaken using principals of motivational interviewing and will be made in weeks 1-12, 14, 16, 18, 21 and 24 for Extended Program. Notepad icons indicate data collection visits.

**Table 1.**
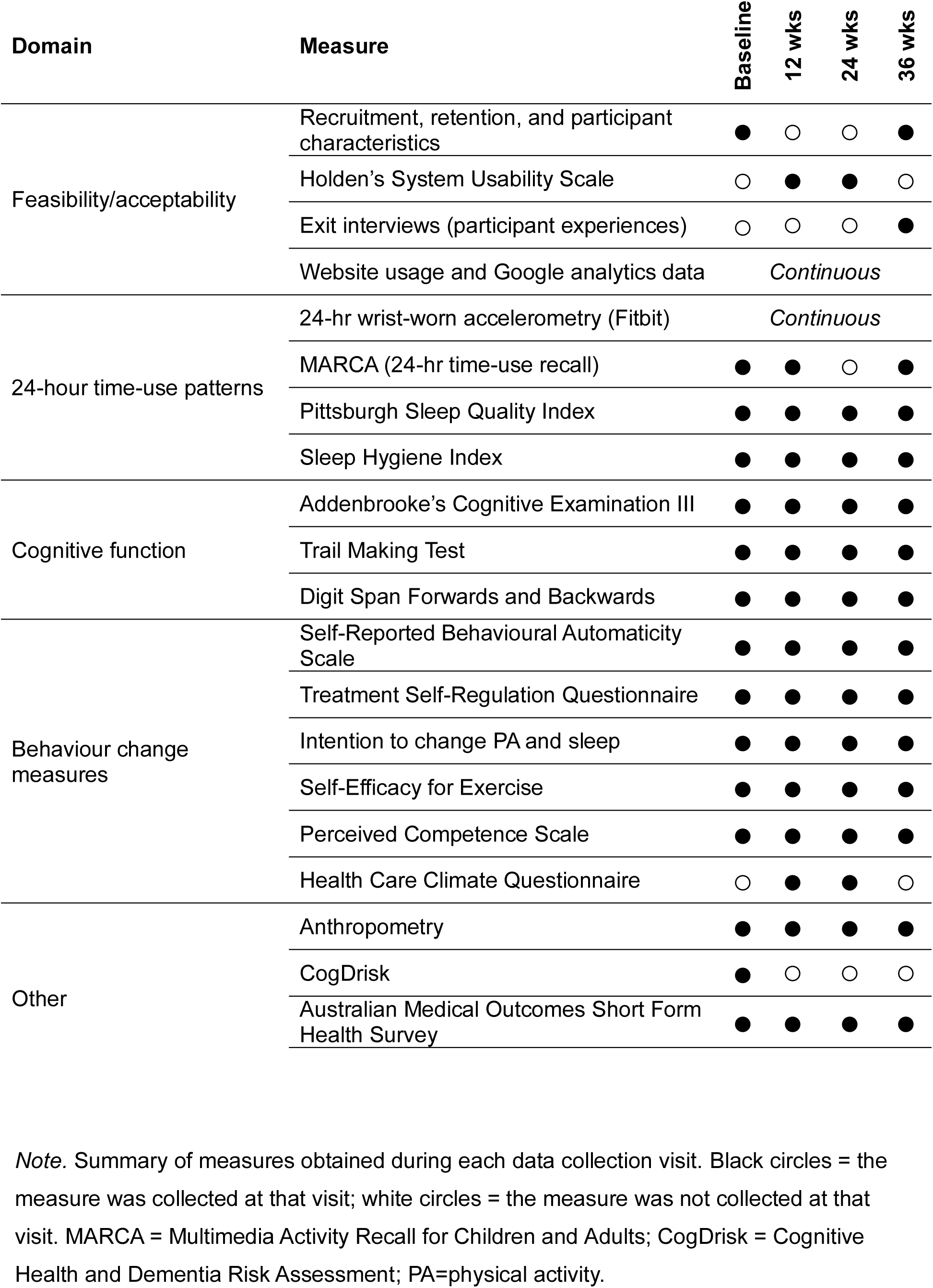
Data collection schedule for primary and secondary outcome measures.

| Domain | Measure | Baseline | 12 wks | 24 wks | 36 wks |
| --- | --- | --- | --- | --- | --- |
| Feasibility/acceptability | Recruitment, retention, and participant characteristics | ● | ○ | ○ | ● |
|  | Holden's System Usability Scale | ○ | ● | ● | ○ |
|  | Exit interviews (participant experiences) | ○ | ○ | ○ | ● |
|  | Website usage and Google analytics data | *Continuous* |  |  |  |
| 24-hour time-use patterns | 24-hr wrist-worn accelerometry (Fitbit) | *Continuous* |  |  |  |
|  | MARCA (24-hr time-use recall) | ● | ● | ○ | ● |
|  | Pittsburgh Sleep Quality Index | ● | ● | ● | ● |
|  | Sleep Hygiene Index | ● | ● | ● | ● |
| Cognitive function | Addenbrooke's Cognitive Examination III | ● | ● | ● | ● |
|  | Trail Making Test | ● | ● | ● | ● |
|  | Digit Span Forwards and Backwards | ● | ● | ● | ● |
| Behaviour change measures | Self-Reported Behavioural Automaticity Scale | ● | ● | ● | ● |
|  | Treatment Self-Regulation Questionnaire | ● | ● | ● | ● |
|  | Intention to change PA and sleep | ● | ● | ● | ● |
|  | Self-Efficacy for Exercise | ● | ● | ● | ● |
|  | Perceived Competence Scale | ● | ● | ● | ● |
|  | Health Care Climate Questionnaire | ○ | ● | ● | ○ |
| Other | Anthropometry | ● | ● | ● | ● |
|  | CogDrisk | ● | ○ | ○ | ○ |
|  | Australian Medical Outcomes Short Form Health Survey | ● | ● | ● | ● |

*Note.* Summary of measures obtained during each data collection visit. Black circles = the measure was collected at that visit; white circles = the measure was not collected at that visit. MARCA = Multimedia Activity Recall for Children and Adults; CogDrisk = Cognitive Health and Dementia Risk Assessment; PA=physical activity.

#### Feasibility and acceptability of Small Steps

The primary outcome measures of the *Small Steps* study are the feasibility and acceptability of the intervention, which will be quantified in several ways. First, we will measure the number and characteristics of participants initially recruited and randomised. Second, we will measure adherence to the intervention protocol as: 1) the number of withdrawals or losses to follow-up during the intervention; 2) the time (weeks) between participant randomisation and withdrawal, and reasons why; 3) the percentage of weekly check-ins completed on the website (of a possible 12 scheduled for the Extended program); 4) the percentage of phone calls completed (of a possible 17 scheduled for the Extended program); and 5) the number of reminders sent to the participant to sync and/or charge their Fitbit device across the intervention period. The impact of intervention adherence will be presented as summary statistics and assessed through sensitivity analyses, rather than quantifying feasibility based on a pre-determined minimum adherence threshold (25). Third, we will examine feasibility, fidelity and enjoyment through analysing responses from the post-intervention semi-structured interviews (detailed below). Finally, we will measure safety of the intervention through reporting of adverse events. Additionally, at week 12 and 24, participants will complete a modified version of Holden’s Simplified System Usability Scale (SUS (26)) to provide feedback on the perceived usability of the *Small Steps* website. Participants will respond to 10 items on a 5-point Likert-type scale ranging from 1 (strongly disagree) to 5 (strongly agree). For the purpose of this study, the term ‘system’ will be replaced with ‘Small Steps website’ (such as ‘*The Small Steps website is too complex for me’*, and ‘*The various parts of the Small Steps website were well integrated’*).

To understand acceptability of the internet-delivered intervention, website usage and engagement will be tracked across several metrics. The *Small Steps* website will gather participant-level data on goals set at the beginning of the intervention and characteristics of check-ins made across the intervention (i.e., type and frequency of goals set). We will supplement this information with the website’s Google Analytics data, by gathering cohort and/or individual-level information on number of visits and time spent on specific pages within the website (e.g., check-ins, resources) to track engagement.

Following their final testing visit (Week 36), participants will be invited to complete a brief semi-structured exit interview with a member of the research team who was not involved in intervention delivery or testing (AS). Interviews will explore participants’ perceptions and expectations of the *Small Steps* intervention with specific question prompts to better understand engagement, barriers, improvements and assessment burden. Interviews will be transcribed verbatim and thematically coded using inductive thematic analysis (27–29). The semi-structured interview guide can be viewed in Supplementary File 4.

### Secondary outcome measures

Secondary outcomes of the *Small Steps* study include: preliminary effectiveness quantified as the change in 24-hour time-use patterns (including duration, context and quality measures) across the course of the intervention period; cognitive function; correlates and measures of behaviour change; anthropometry; dementia risk factor profile; and quality of life. These measures may be used as study outcomes (e.g., the impact of the intervention on cognitive function), or as predictors in analyses (e.g., assessing whether changes in anthropometric measures (e.g., waist-hip ratio) moderate changes in cognitive function).

#### Twenty-four-hour time-use patterns

Preliminary intervention effectiveness will be measured as the change in device-measured 24-hour time-use patterns. Across the entire study duration, daily time spent in sleep, sedentary behaviour, light intensity PA and moderate-vigorous PA will be measured using continuous wrist-worn accelerometry (Fitbit Inspire 3: Fitbit Inc., San Francisco, CA, USA). Participants will be asked to wear a Fitbit on their non-dominant wrist at all times throughout the study (except for bathing and when charging the device), and will be required to sync their Fitbit data to their Fitbit account (via the Fitbit app) once every 2 days to ensure data retention. Fitbit data collected for this study will include 24-hour time-use patterns (categorised as time in sleep, sedentary behaviour (including awake in bed), light and moderate-vigorous PA across the 24-hr day according to Fitbit’s proprietary algorithm), heart rate, and device settings (name, battery level, last sync). These data will be collected and stored remotely by ‘Fitnesslink’, an in-house software developed by researchers at the University of South Australia and Portal Australia (Adelaide, Australia). The Fitnesslink dashboard will be monitored by research staff to ensure participants (across both conditions) are achieving sufficient wear time, maintaining charge and syncing their data regularly during the intervention.

24-hour time use will also be captured *via* self-report using the Multimedia Activity Recall for Children and Adults (MARCA) tool (30). The MARCA will be conducted *via* telephone call with a trained researcher and requires participants to recall every activity that they engaged in over the past two days from midnight to midnight, in minimum 5-minute increments. The MARCA tool contains over 500 activities linked to an activity compendium, allowing 24-hour time use to be conceptualised in terms of both activity types and activity intensities (through linking each individual activity with its metabolic equivalent per the compendium).

Sleep quality will be collected using the Pittsburgh Sleep Quality Index (PSQI (31)), a self-report questionnaire capturing sleep quality across seven sub-domains (each with their own sub-score) including subjective sleep quality, sleep latency, sleep duration, habitual sleep efficiency, sleep disturbances, use of sleeping medication, and daytime dysfunction. A total score greater than 5 indicates poor sleep quality (31).

Sleep hygiene behaviours will be collected using the Sleep Hygiene Index (32), a 13-item self-report measure where higher scores indicate poorer sleep hygiene behaviour. Items such as “*I take daytime naps lasting two or more hours”* and “*I use alcohol, tobacco, or caffeine within 4 hours of going to bed or after going to bed”* are answered using a five-point Likert-type scale, ranging from ‘never’ to ‘always’.

#### Cognitive function

A brief battery of cognitive assessments will be administered by trained study personnel in a quiet, temperature-controlled room, including the Addenbrooke’s Cognitive Examination III (ACE-III), Trail Making Test (A and B), and Digit Span (forwards and backwards).

The ACE-III (33) is a brief paper-and-pencil-style cognitive test used to assess cognitive function across five domains including memory, attention/orientation, fluency, language, and visuospatial ability. The ACE-III is scored out of 100, with a cut-off score of 88 demonstrating high sensitivity and specificity to detecting dementia in previous studies (33). The version of ACE-III will be alternated at each timepoint to minimise practice effects.

The Trail Making Test (34) is a paper-and-pencil-style cognitive test with two parts, A and B. Part A of the test requires participants to use a pencil to connect a series of 25 encircled numbers (on paper) in numerical order (from 1 to 2 to 3, and so on) as fast and accurately as possible. Part B of the test follows the same principles but follows an alphanumeric pattern whereby the participant must connect 25 encircled numbers and letters in logical order (e.g., from 1 to A to 2 to B, and so on). Part A is widely recognised as a measure of processing speed, whilst Part B is considered a measure of executive function due to the requirement to concurrently manage two streams of information. The time to completion (in seconds) for Parts A and B will be used as outcome measures for these tests.

The Digit Span task (35) is a measure of verbal working memory, which has been shown to decline with increasing age. The task consists of two parts, namely Digit Span Forwards and Digit Span Backwards. During each part, participants listen to a series of digit spans read aloud by the researcher (at a rate of one number per second), and are asked to repeat the digit span back to the researcher in the same order as was read (Forwards), or in the reverse order (Backwards). Digit spans range in length from three to nine digits in part one (Forwards), two to eight digits in part two (Backwards), and two trials are performed for each digit span. The maximum number of digits correctly repeated without error (in either of the two trials) are calculated for both Forwards and Backwards conditions, and thus each participant will have two scores for the Digit Span task.

To optimise their sensitivity to change over time, researchers will aim to undertake cognitive assessments in similar conditions (e.g., time of day, in a temperature-controlled room) and in the same order at each timepoint.

#### Measures of behaviour change

Several self-report measures will be used to assess changes in perceived behavioural automaticity, self-efficacy for PA, intention to change PA and sleep, motivation, competence, and autonomy support, which are known predictors of behaviour change.

The Self-Report Behavioural Automaticity Index (SRBAI) (36) will be administered to assess habit strength in relation to PA, sleep, and sedentary behaviours. Participants will be asked to rate four items (such as “*Physical activity is something…1) I do automatically, 2) I do without having to consciously remember, 3) I do without thinking, and 4) I start doing before realising I’m doing it”*) on a scale ranging from 1 (strongly disagree) to 7 (strongly agree).

Participants will complete the four items for each PA, sedentary behaviour, and good sleep practices (12 items total). Participants will be provided with a definition of each of these three concepts prior to rating the four items.

Participants’ motivation to be physically active will be captured using the Treatment Self-Regulation Questionnaire (37, 38). Participants will respond to 15 questions relating to reasons for beginning or keeping regular disease self-management, such as “*Because I personally think it is best for my health”* and “*Because I would feel guilty or ashamed if I did not manage my own disease”* using a 7-point Likert-type scale (1=not true at all, 7=completely true).

Intention to change PA and sleep will be measured using two statements: ‘I intend to engage in physical activity, at least every other day, over the next 3 months’; and ‘I intend to follow good sleep habits over the next 3 months’. Participants are asked to indicate their alignment with each statement, ranging from 1 (strongly disagree) to 7 (strongly agree).

Self-efficacy for PA will be measured using the Self-Efficacy for Exercise Scale (39). For this measure, participants will respond to the question “*How confident are you right now that you could exercise 3 times per week for 20 minutes if…”* across nine different statements, such as “*The weather was bothering you”*, or “*You felt stressed”*. Responses range from 0 (not very confident) to 10 (very confident) on an 11-point Likert-type scale.

Perceived competence to exercise will be captured using an adapted Perceived Competence Scale (40). Participants will respond to four items relating to their confidence and perceived capability to engage in PA (e.g., “*I now feel capable of being physically active regularly”* on a 7-point Likert-type scale, ranging from 1 (not at all true) to 7 (very true).

To assess participants’ perception of the autonomy support provided by the *Small Steps* health professional (i.e., the trained researcher administering the intervention), we will administer the Health Care Climate Questionnaire (HCCQ) (40). Participants will respond to six items (e.g., “*I feel that my Small Steps health professional has provided me with choices and options about changing my physical activity and sleep behaviour (including not changing)*” using a 7-point Likert-type scale, ranging from 1 (not true at all) to 7 (very true).

#### Anthropometry

Height (cm; measured with portable Leicester Height Measure), weight (kg; measured with Withings Body scales), waist and hip circumference (cm; measured with Lufkin tape measure) will be measured using standardised protocols according to the International Standards for Anthropometric Assessment (41). Each will be measured twice, and the average of the two measures will be calculated and used as the outcome. Additionally, we will calculate body mass index (BMI; body mass (kg) divided by height-squared (m^2^)) and waist-hip ratio (waist girth divided by hip girth).

Resting blood pressure (both systolic and diastolic) will be assessed using a digital blood pressure monitor (OMRON Healthcare Co. model 1A1B Hem-7000-CIL) placed on the upper non-dominant arm (approximately 2-3cm above the anti-cubital fossa). Three measures will be taken at least one minute apart, and average values for each systolic blood pressure, diastolic blood pressure and resting heart rate calculated.

#### Dementia risk profile

At baseline, participants will complete the Cognitive Health and Dementia Risk Assessment (CogDrisk (42)) to capture exposure to modifiable and non-modifiable risk factors for dementia. The CogDrisk consists of ∼90 questions that address 17 risk factors for dementia including age, sex, education, hypertension, midlife high cholesterol, midlife obesity, diabetes, insufficient PA, depression, traumatic brain injury, atrial fibrillation without stroke, smoking, social engagement, cognitive engagement, diet, pesticide exposure, stroke, and insomnia.

#### Health-related quality of life

Health-related quality of life will be measured using the Medical Outcomes Study Short Form Health Survey (SF-12v2; (43)). The 12-item survey assesses perceived overall health and dimensions of physical health (e.g., physical functioning, bodily pain) and mental health (e.g., vitality, social functioning). Perceived physical and mental health will be identified through generating physical and mental component summary scores using standardised scoring instructions.

#### Demographic covariates

Demographic information including age, sex, highest educational attainment, retirement status, current/previous occupation and marital status will be collected at baseline using REDCap data management software (24).

### Small Steps website

The *Small Steps* intervention will be delivered *via* a custom website. Prior to this RCT, the website was created through a co-design process involving research staff, web developers, community-dwelling older adults and health professionals (e.g., clinical exercise physiologists), described in detail elsewhere (20). The utility of the website throughout the intervention is described in the following sections.

### Study procedure

The intervention will involve three main phases, the Introductory, Maintenance, and Follow-up phases, each lasting 12 weeks (Figure 1). Each of these phases is followed by an in-person data collection visit as described previously (see Table 1 for list of measures collected at each visit).

At the beginning of *Small Steps* all participants will complete their baseline data collection visit. First, following confirmation that participants understand and are comfortable with study procedures and expectations, written informed consent will be obtained. During this initial visit, participants will also be given their Fitbit watch to wear continuously for the duration of the study. To ensure that at least 7-days of 24-hr time-use data have been collected by the Fitbit, participants will return for their ‘onboarding’ visit at least one week following the baseline visit. Between baseline and onboarding visits, participants will also complete their first MARCA phone call with a research staff member. Step count (from Fitbit data) along with other variables previously outlined (‘Participant randomisation and blinding protocol’ section) will be used to randomise the participants to either the Extended or Condensed (i.e., intervention or control) program. Protocol for the Extended and Condensed conditions are described separately below.

#### Extended program protocol

##### Onboarding and goal setting

Onboarding visits will be conducted in-person with a member of research staff (BR). The main component of the onboarding visit will be to set personalised behaviour change goals with the participant. Participants will firstly be asked to identify an ‘overall’ personal outcome goal for the *Small Steps* program, such as “to be more active to keep up with the grandchildren”. This overall outcome goal will feature on the *Small Steps* website dashboard (Figure 2) throughout the intervention to promote ownership and intrinsic motivation.

Additionally, participants will be asked to set a specific behaviour change goal, one for PA (e.g., “replace 30 minutes of sitting time per day with 30 minutes of physical activity”) and an additional optional goal for sleep, which they will aim to achieve by the end of the Introductory phase. These specific goals will be set using a SMART (Specific, Measurable, Attainable/Achievable, Relevant/Realistic, Timed) goals framework (44). A step-by-step demonstration of the Onboarding protocol is provided in Supplementary File 1.

Several tools will be used to assist the participant in setting their overall behaviour change goal. The primary tool will be a comparison of their current 24-hour time-use composition (collected by Fitbit) to their ‘ideal’ 24-hour time-use composition. The ‘ideal’ 24-hour day for each participant will be generated using compositional optimisation analyses (45) performed on the UK Biobank dataset (46), personalised to the individual’s sociodemographic characteristics and exposure to dementia risk factors. The statistical methods and co-designed interactive interface used to create and display the ideal day will be described in detail in a future manuscript. In addition to the ‘ideal day’ recommendation generated for the participant, where required, the researcher will also display data collected by the MARCA using a custom app developed in R Shiny (Supplementary File 2), as well as information about their sleep quality and hygiene collected using the PSQI and Sleep Hygiene Index (as needed). These measures will aim to provide further insight into the types and contexts of daily activities, or components of sleep hygiene, which could be targeted in their intervention.

With support from the researcher, the participant will then identify behaviours that they may wish to try to reduce or replace (e.g., taking the elevator) and behaviours that they may want to implement or increase (e.g., taking the stairs) within the Introductory phase to achieve their goal. Participants will have access to a pre-filled list of behaviours to choose from but will also have autonomy to add their own. These selections will be saved against the participant’s profile and appear as behaviour change options during their weekly check-ins (described below).

Finally, the researcher will familiarise the participants with the *Small Steps* website and will assist them to complete their week 1 ‘check-in’, detailed in the following section.

#### Introductory phase

During the 12 weeks of the Introductory phase, participants will independently complete weekly ‘check-ins’ using the *Small Steps* website. Weekly check-ins will require participants to make behaviour change choices (i.e., action plans) which move them closer to their goal for the *Small Steps* intervention. At the beginning of each check-in (except Week 1), participants will firstly self-report the success of the preceding week’s action plan on a 5-point Goal Attainment Scale, rating from ‘much worse than expected’ to ‘much better than expected’. Participants will then make an action plan (targeting either PA or sleep) for the week ahead. To promote autonomy, participants’ weekly choices will be customisable.

Participants will identify the behaviour they want to replace, the new behaviour they want to implement, as well as the duration (for PA-related choices) and/or weekly frequency of the behaviour, when/where the new behaviour will be completed, with whom the behaviour will be completed (PA only), and will be presented with a free-text box with the prompt “what do you need to achieve this?” (e.g., “I will write a reminder on the fridge”). Supplementary File 3 displays an example check-in. Based on participant feedback in the co-design and beta testing of *Small Steps*, an additional option of “no new goal” was added if the participant did not want to set a new action plan for that week (e.g., as a result of not achieving the previous goal).

Weekly supportive phone calls conducted using the principles of motivational interviewing (∼5-10 minutes each) will be made by a *Small Steps* researcher throughout the Introductory phase. During the phone calls, the researcher will ask the participant to briefly reflect on their progress, and to discuss barriers and challenges in achieving their goal (where required).

Any adverse events (whether related to the intervention or not) will be recorded during this phone call and reported to ethics. Additionally, during the phone calls in Weeks 4, 8, and 12, the researcher will provide feedback on current 24-hour time-use patterns (i.e., time spent in PA, sleep, and sedentary behaviour, averaged over the previous 7 days) from Fitbit monitoring.

For the duration of the intervention (from Introductory phase to completion of Follow-up phase), participants will have access to educational resources *via* the website (Resources tab on home page). Educational material obtained from existing credible sources (47–50) includes general information about the benefits of being physically active, relationships between physical activity, sleep and dementia, as well as more specific resources containing strategies to improve physical activity and sleep. Three home-based exercise plans (each with two levels of difficulty) created by the research team will be made available to participants, covering stretching, balance and strength exercises. Participants in the Extended program will have access to an additional 5 resources outlining available physical activity programs in their local area, including the cost of the programs. Following each weekly check-in, one resource relating to the type of choice made is displayed on the website dashboard.

#### Maintenance phase

During the 12-week Maintenance phase, participants will be instructed to at least maintain, or continue improving, 24-hour time-use patterns implemented during the Introductory phase. Participants will not be required to make weekly check-ins, but will continue to receive regular supportive phone calls from the researcher. The frequency of phone calls will reduce over the course of the Maintenance phase, occurring fortnightly for the first 6 weeks (Weeks 14, 16, and 18), and every 3 weeks thereafter (Week 21 and 24). Fitbit 7-day averages will be discussed during these calls to update participants on their current time use. Participants will continue to have access to the website to see previously set goals, their overall goal for the intervention, and educational resources.

#### Follow-up phase

The final phase of *Small Steps* is a 12-week Follow-up phase, during which participants will be encouraged to maintain the 24-hour time-use improvement strategies implemented during the previous phases. Participants will not receive any phone calls from the researchers during Follow-up.

#### Condensed program

Participants allocated to the Condensed (control) program will attend an initial Onboarding session which will follow the same goal-setting protocol as those in the Extended (intervention) program. Participants will be educated on the functionality of the *Small Steps* website, which will be restricted with access to the educational resources only. Weekly check-ins, tailored resource provision and supportive phone calls will not be provided to participants in this group. The researcher will inform the participants that they will not have any contact with research staff for the remainder of the program, except during data collection sessions at baseline, 12, 24 and 36-week timepoints, or where research staff are required to remind the participant to charge or sync their Fitbit.

### Mapping of *Small Steps* to Behaviour Change Theory (BCT)

Few multi-domain dementia risk-reduction interventions code intervention elements to established behaviour change frameworks such as the Behaviour Change Technique Ontology (BCTO) (51) or the Behaviour Change Taxonomy (BCTT) (52), limiting ability to compare the building blocks of behaviour change between interventions. The intervention elements of *Small Steps* were mapped to the 20 higher-level groups of the BCTO and are presented in Table 2.

**Table 2.**
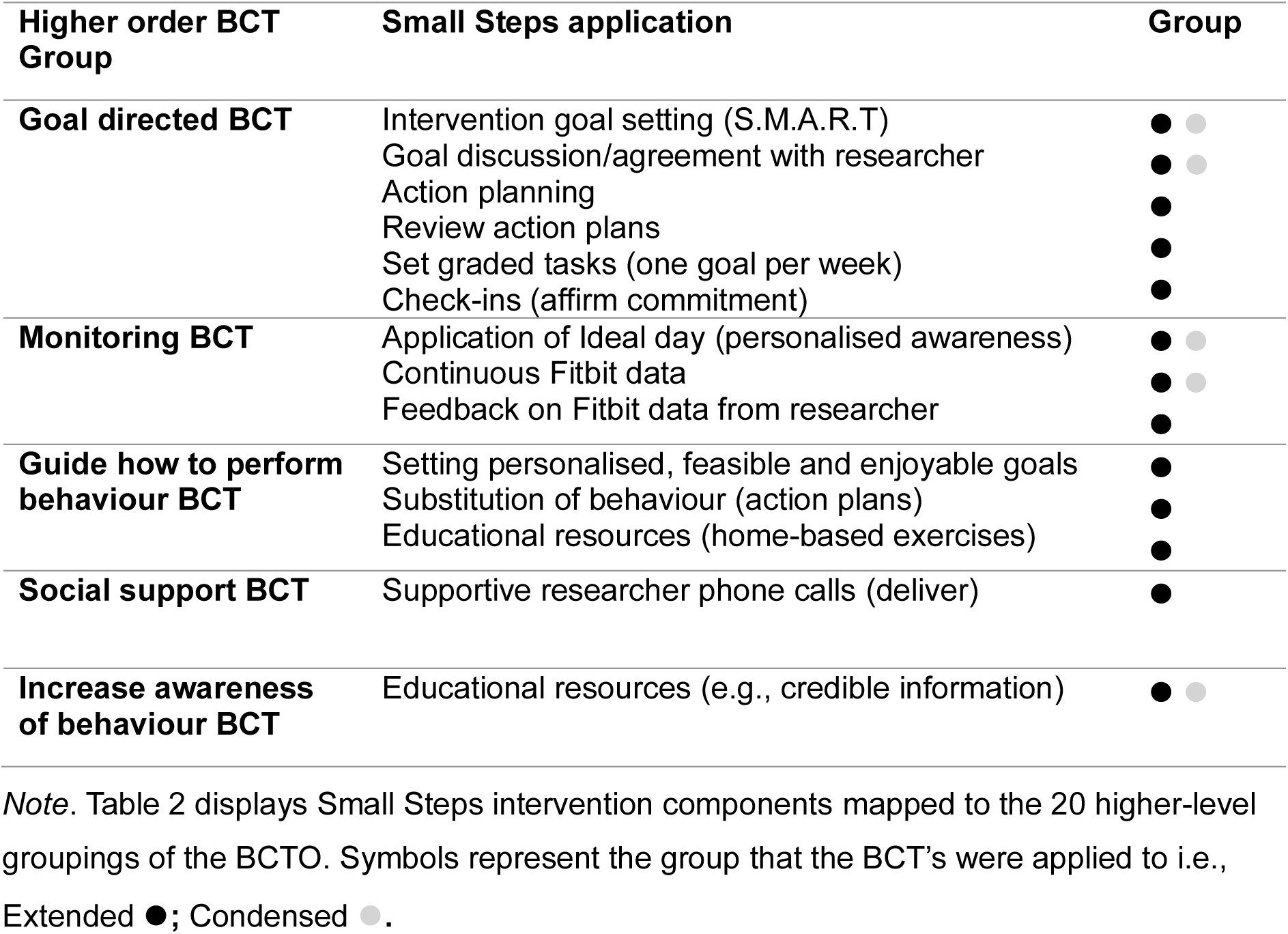
Behaviour Change Techniques applied in Small Steps intervention.

### Data analysis

#### Feasibility and acceptability of *Small Steps*

Upon completion of the study a CONSORT diagram will be generated to display participant retention, and where necessary, reasons for withdrawal at each intervention phase.

Engagement with the intervention will be described using website and Google Analytics metrics. We plan to conduct analyses to determine the influence of adherence on intervention effectiveness, using continuous measures (%) of adherence described previously. Post-intervention exit interviews will be thematically coded using inductive thematic analysis to explore facilitators and barriers to engagement with the intervention, and to obtain suggestions for improvement.

#### Preliminary intervention effectiveness

For each data collection visit (Baseline, Week 12, Week 24 and Week 36) and group (Extended and Condensed), we will present descriptive statistics detailing each outcome measure including time-use characteristics (mean time spent in sleep, sedentary behaviour, LPA and MVPA in the week prior to the visit), anthropometric measures, cognitive test scores, and intervention engagement (number of check-ins within each phase, for Extended program only). Categorical variables will be presented as proportions, and continuous variables reported as means (with standard deviations) for symmetric distributions, or medians (with interquartile range) for asymmetrically distributed data. Missingness will be assessed prior to analyses. If the proportion of participants with a missing data collection visit (one or more) is greater than 10% then multilevel imputation will be used, otherwise complete case analysis will be undertaken. All tests will be two-tailed and use α=0.05 to determine significance.

A linear mixed-effects model will be used to regress 24-hr time-use composition as a multivariate outcome on intervention group (Extended or Condensed program), timepoint (Baseline, Week 12, Week 24, Week 36), and a time*group interaction as fixed effects to examine group over time differences in 24-hr time use. Analyses will be adjusted for baseline demographic covariates of age, sex, education, retirement status, and marital status.

Random intercepts and slopes (if warranted by residual diagnostics, the parameterisation of time, and reduction in Bayesian Information Criterion goodness-of-fit statistic) will account for repeated measures on participants. Post-hoc contrasts between groups at each follow-up data collection time-point (with 95% confidence intervals) will be calculated from the linear mixed-effect model fit to estimate average 24-hr time-use composition change from Baseline (difference of differences) upon back-transformation to the time-use composition scale. The secondary outcome of change in cognitive function from baseline (as a univariate response) will be regressed on baseline cognitive function, group, timepoint, and time*group interaction (fixed) with adjustment for demographic covariates (fixed). Random intercepts for participant will account for repeated measures over time. Additionally, we will also undertake exploratory analysis to determine whether behaviour change variables (i.e., motivation-related outcome measures, collected at baseline and follow-up timepoints) moderated any group and time-use associations.

### Ethics and dissemination

#### Data management and dissemination

Data collected during testing at all four timepoints and during the Introductory and Maintenance phases (phone calls) will be managed using REDCap (24), an electronic data management application hosted at the University of South Australia. Where data files exceed size requirements for REDCap we will store them in a secure online cloud storage platform (Nextcloud, also hosted by the University of South Australia). Continuously collected Fitbit data will be harvested and stored by Fitnesslink on Amazon Web Services servers located within Australia. Fitnesslink data will be exported and backed up on Nextcloud regularly throughout the study. The *Small Steps* website, which will store weekly check-in data inputted by the participants, was developed within WordPress and is hosted onshore by VentraIP who partner with NEXTDC for data storage. All four data storage platforms (REDCap, Nextcloud, Fitnesslink and the *Small Steps* website) will require a username and password to access the data, which will only be provided to authorised project team members. Data will be stored for 15 years following study completion, as required by ethics. Only investigators who are listed on ethics approvals will have access to the data, and access rights will be managed by the Chief Investigator (AS). Due to the small sample size and low-risk nature of the trial, an external data monitoring committee will not be formed.

Rather, data quality (e.g., completeness) and integrity will be frequently monitored by trial staff (CY, EW, BR) and any detected issues will be raised with the wider authorship team for action.

It is anticipated that the findings of *Small Steps* will contribute to numerous peer-reviewed publications, and will be presented at both national and international conferences. Data will be published once all participants (n=88) have completed final data collection at Week 36.

#### Potential risks and risk management

All participants will be provided with detailed information about all components of the study prior to providing written informed consent. Participants will be informed that they can elect to withdraw from any individual test (e.g., cognitive testing) or the wider study at any stage. Research staff will also confirm with participants whether they wish to withdraw use of any data leading up to their withdrawal. In the case that participants become distressed during the study, staff will follow a participant distress protocol and ensure that additional supports are offered and/or referred to, where appropriate.

Participating in any form of physical activity may carry some risk of pain, discomfort, or injury. We have sought to minimise these risks through thorough screening of participant’s conditions and potential contraindications to exercise types, and through ensuring participants are adequately briefed on safe exercising and gradual progression in intensity or load.

### Data availability

Following completion of the *Small Steps* study and publication of all research articles identified by the research team, de-identified data will be available to other research groups upon reasonable request. To obtain an ethically compliant data set (with de-identified data), researchers will contact the corresponding author and provide evidence of institutional ethics approval for local data storage, analysis and security.

## DISCUSSION

The *Small Steps* study will be the first of its kind to examine the feasibility, acceptability and preliminary effectiveness of a co-designed personalised behavioural intervention to optimise 24-hour time-use composition (i.e., the balance of physical activity, sleep and sitting in the 24-hour day) for cognitive health in older adults without dementia. We expect that the personalised and flexible nature of the intervention will promote autonomy and result in sustained behaviour change, and that it will be acceptable to the study population due to it having been co-designed with members of the target population group. In addition to changes in behaviour across the intervention, we will collect regular measures of perceived personal autonomy and motivation of the participant, as well as their experience with using the website as an intervention tool. The secondary outcomes of the intervention, including cognitive function measures, will provide important insight into the potential of personalised 24-hour time-use interventions to reduce the risk of dementia. If deemed acceptable and feasible, further funding will be sought to scale this up.

## ACKNOWLEDGEMENTS

We acknowledge the significant contributions of City of Onkaparinga and ACH Group as study partners and thank them for their support in all phases of *Small Steps*. We thank Redshift Creative for their collaboration as software developers of the *Small Steps* website.

## AUTHOR CONTRIBUTIONS

MLM, HTB, AD, and AES ran co-design workshops to conceptualise the study. KL and AES ran initial community consultation to inform study design. HTB, BR and AES developed the website. MLM, DD, TS and AES developed the behaviour change app. MLM, TF, and AES developed the accelerometry data collection protocol. MLM, HTB, HADK, AMC, ATW, ET, CY and AES developed primary and secondary measures. MLM, DD, TO, TS, and AES developed the statistical approach. MLM, HTB, BR and AES prepared the manuscript. All authors reviewed the manuscript and approved the final version.

