## Supplementary File for "‘Small Steps’ towards improving 24-hour time-use behaviours to decrease the risk of dementia: protocol for a personalised, web-based randomised controlled trial in community-dwelling older adults"

**Supplementary File 1.** Goal-setting and participant onboarding

The following screenshots display the onboarding process that is undertaken by the participant, with support from a research staff member, at the beginning of the Small Steps intervention. Onboarding is undertaken on the Small Steps website while logged in to the participant's account.

**Step 1: *Select the focus of the Small Steps program*.**

Participants are able to select 'Be more physically active', or 'Improve my sleep health', or both as their focus.

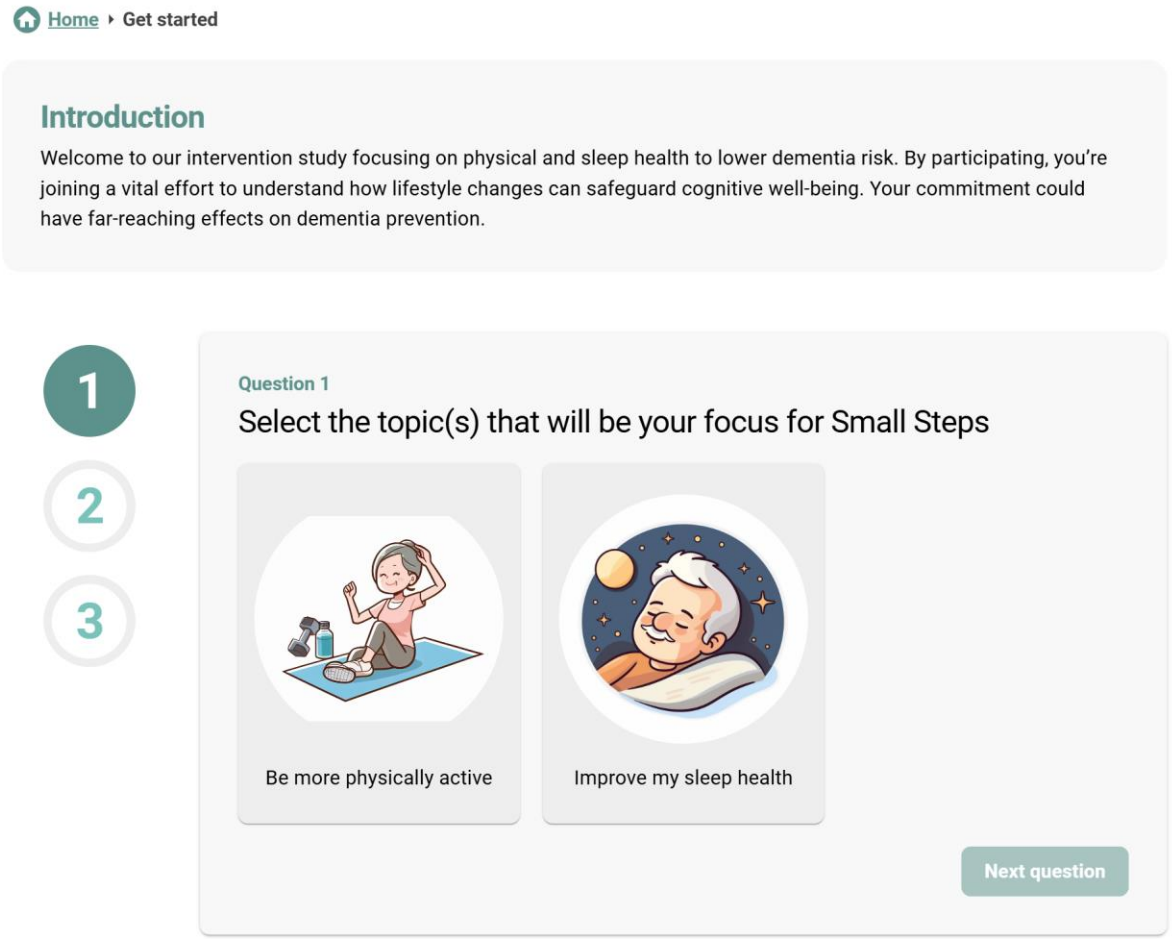

**Step 2:** ***Set overall and specific goals for the program.***

Participants are firstly asked to write a small description of their overall goal for the Small Steps program. An example may be ‘I want to maintain my mobility so that I can keep up with the grandkids’. Then, following the SMART goal approach, participants are asked to outline a more specific physical activity and/or sleep-related goal. For example: ‘By the end of this program I will have increased my physical activity by 20 minutes per day, and will take that time away from watching TV’.

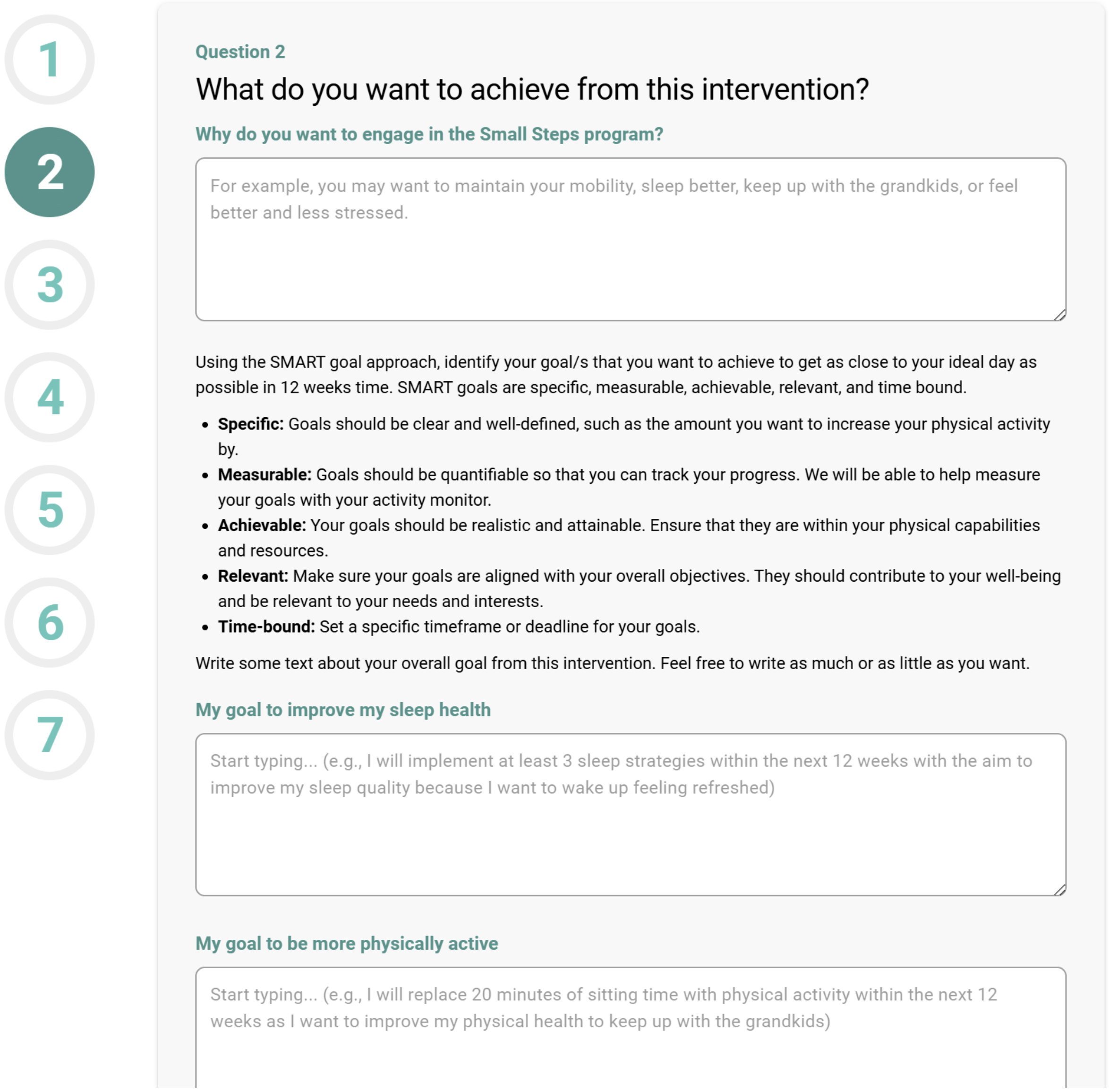
1
2
3
4
5
6
7

Question 2

What do you want to achieve from this intervention?

Why do you want to engage in the Small Steps program?

For example, you may want to maintain your mobility, sleep better, keep up with the grandkids, or feel better and less stressed.

Using the SMART goal approach, identify your goal/s that you want to achieve to get as close to your ideal day as possible in 12 weeks time. SMART goals are specific, measurable, achievable, relevant, and time bound.

- **Specific:** Goals should be clear and well-defined, such as the amount you want to increase your physical activity by.
- **Measurable:** Goals should be quantifiable so that you can track your progress. We will be able to help measure your goals with your activity monitor.
- **Achievable:** Your goals should be realistic and attainable. Ensure that they are within your physical capabilities and resources.
- **Relevant:** Make sure your goals are aligned with your overall objectives. They should contribute to your well-being and be relevant to your needs and interests.
- **Time-bound:** Set a specific timeframe or deadline for your goals.

Write some text about your overall goal from this intervention. Feel free to write as much or as little as you want.

My goal to improve my sleep health

Start typing... (e.g., I will implement at least 3 sleep strategies within the next 12 weeks with the aim to improve my sleep quality because I want to wake up feeling refreshed)

My goal to be more physically active

Start typing... (e.g., I will replace 20 minutes of sitting time with physical activity within the next 12 weeks as I want to improve my physical health to keep up with the grandkids)

**Step 3.** ***Identify current sedentary behaviours that could be reduced as part of the program.***

To enable more personalised goal setting, participants are asked to identify sedentary behaviours that they currently engage in which could be replaced as part of their goals across the duration of the program. Participants can use pre-selected behaviours from the list, or can add custom sedentary behaviours which will then appear in their list (in these screenshots, the custom behaviour is labelled 'user example').

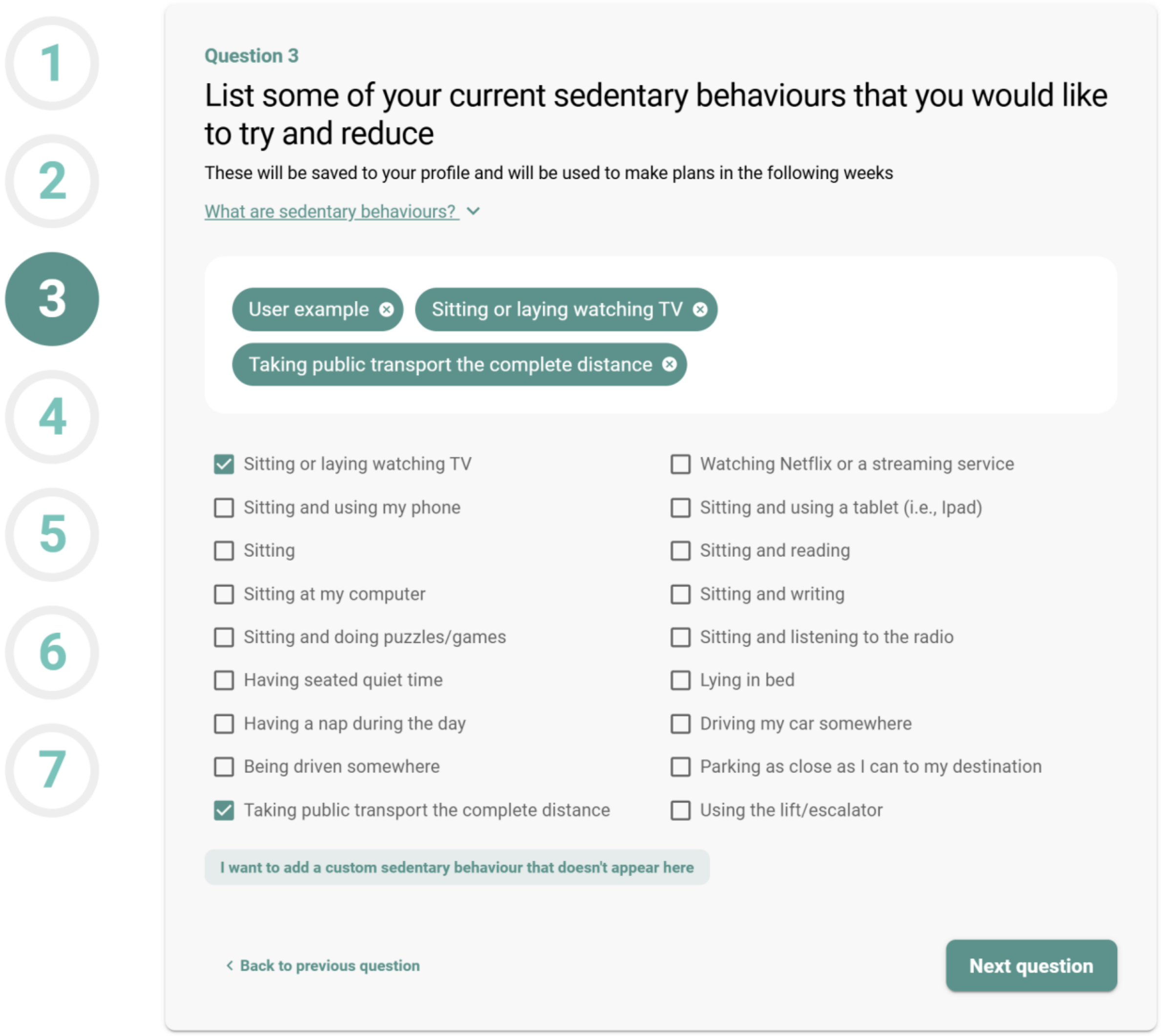

**Step 4.** ***Identify physical activities that could be introduced or increased as part of the program.***

Participants are then asked to identify physical activities that they could introduce or increase during the program. Participants can use pre-selected behaviours from the list, or can add custom physical activities which will then appear in their list (in these screenshots, the custom behaviour is labelled ‘user example’). To help participants, the list of previously selected sedentary behaviours are displayed at the top of the screen. This aims to help participants identify physical activities that could feasibly replace the specific sedentary behaviours they are currently engaging in (i.e., it may not be feasible to replace ‘taking public transport the complete distance’ with ‘taking the stairs’ – rather, this should be replaced with ‘getting off public transport a stop/station early and walking’).

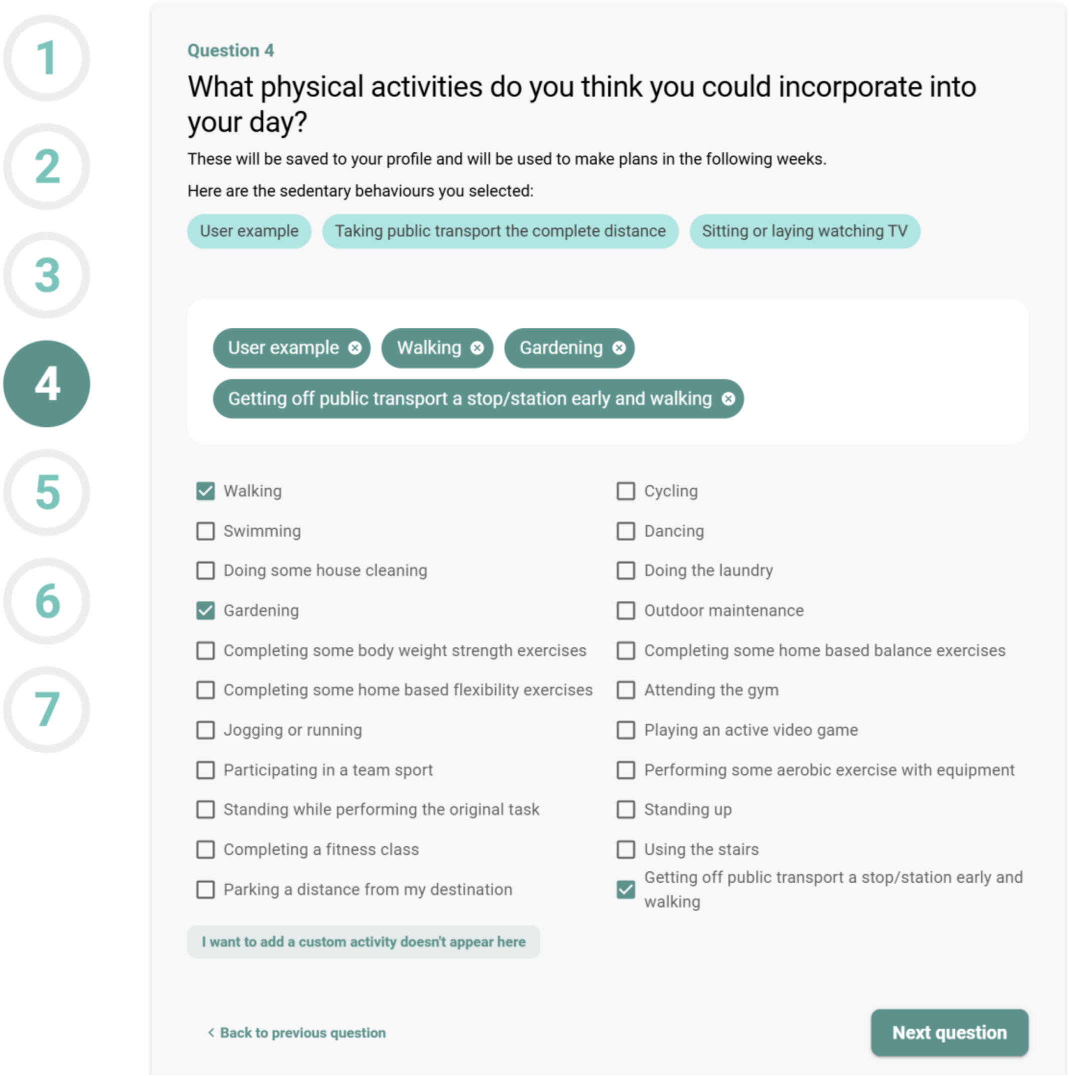

**Step 5.** ***Identify sleep behaviours which may be impairing sleep currently.***

Participants who selected 'improve my sleep health' as one of their overall goals in Step 1 will then be prompted to identify behaviours that may be impairing their sleep currently, which they could target during the Small Steps program. Participants can use pre-selected behaviours from the list, or can add custom sleep behaviours which will then appear in their list.

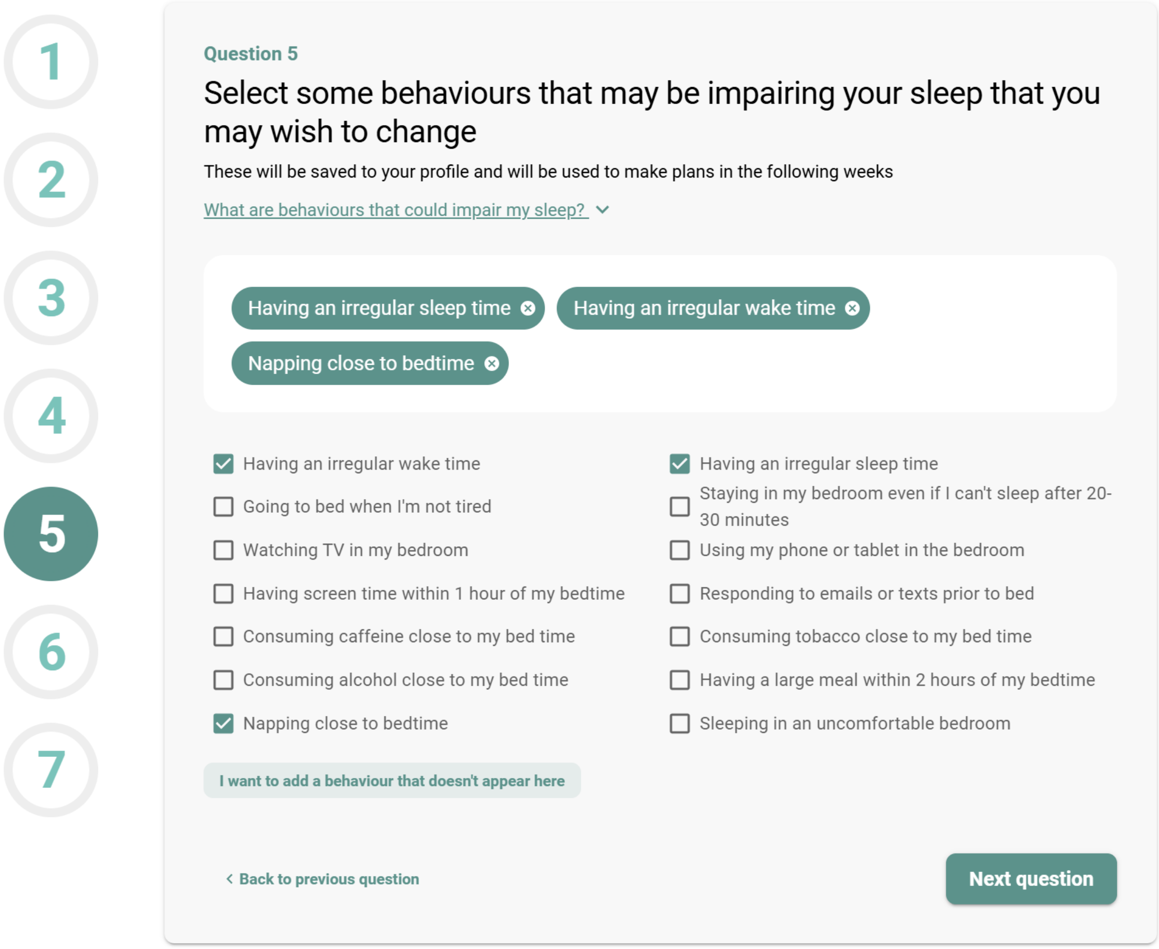

**Step 6.** ***Identify sleep behaviours that could be introduced or increased as part of the program.***

Participants are then asked to identify sleep behaviours that they could introduce or increase during the program. Participants can use pre-selected behaviours from the list, or can add custom sleep behaviours which will then appear in their list. To help participants, the list of previously selected behaviours which are negatively impacting their sleep are displayed at the top of the screen. This aims to help participants identify sleep behaviours that could feasibly replace the specific behaviours they are currently engaging in (i.e., it may not be feasible to replace 'having an irregular wake time' with 'doing a crossword – rather, this should be replaced with 'having a strict wake up time').

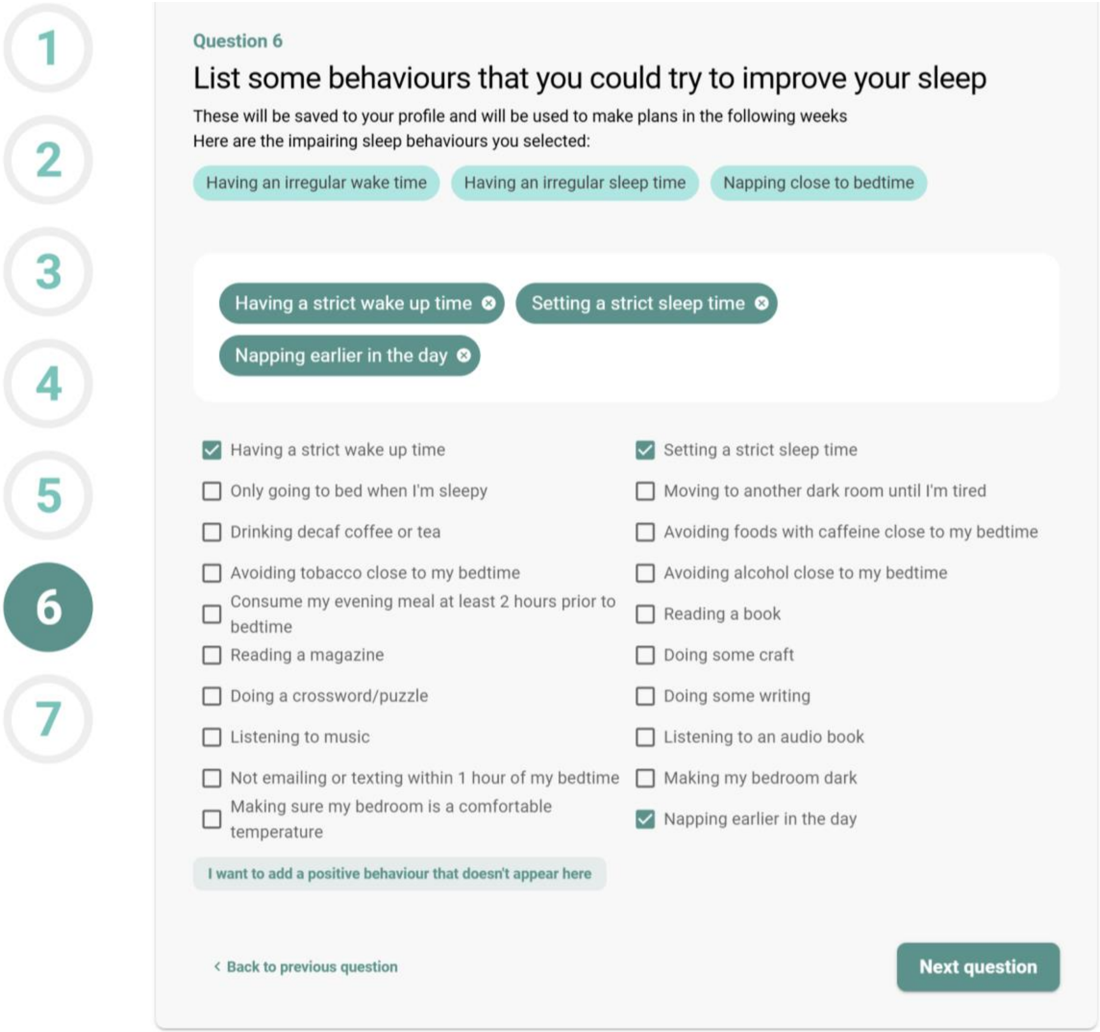

**Step 7.** ***Onboarding is now complete.***

All responses from Steps 1-6 are saved against the participant's Small Steps website account. Overall goals for the intervention (Step 2) will be displayed on the website dashboard under their progress bar (below; 'User example'). The behaviour change options selected in Steps 3-6 will be displayed during weekly check-ins (Supplementary File 3).

### Welcome back, User!

You are now in Week 1 of the initial 12-week Small Steps program. Keep up the great work.

**Week 1** **Week 6** **Week 12**

A reminder of why you are completing Small Steps: User example.

**Click here to complete your next check-in!**

### User, it's time to set some goals!

#### What happens next?

You've completed the onboarding module - and we really appreciate you taking the time to work through it. The next step is to complete the **Week 1 module**, which will allow you to actually get started with deciding on a new activity to implement into your day-to-day life!

**Start Week 1**

**Not ready to start yet?**

Don't worry - you can always come back here later. Just remember to bookmark this page so you can easily find it next time.

Tip: Press **Ctrl + D** on your keyboard to bookmark this page, or look for a ★ bookmark icon on your device's menu

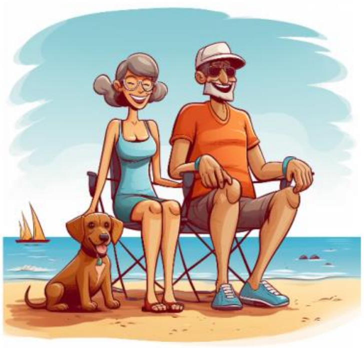

**Supplementary File 2.** MARCA visualisation tool used during Onboarding

The screenshot below displays an example of a Small Steps participant's 2-day recall using the Multimedia Activity Recall for Children and Adults (MARCA) tool. During onboarding, participants are shown their recalled time use by 'superdomain' (right-hand figure legend). This allows the researcher to highlight parts of the day where participants may wish to change their time use as part of the intervention. For example, the example participant below may wish to break up their screen time in the evenings.

Horizonal dotted lines represent threshold whereby the intensity of activity changes. Behaviours below the dotted line at 1.5 METs (y-axis) are sedentary (excluding sleep); behaviours between the two dotted lines (1.5-3 METs) are light intensity physical activities; activities above the dotted line at 3 METs are moderate-to-vigorous intensity physical activities.

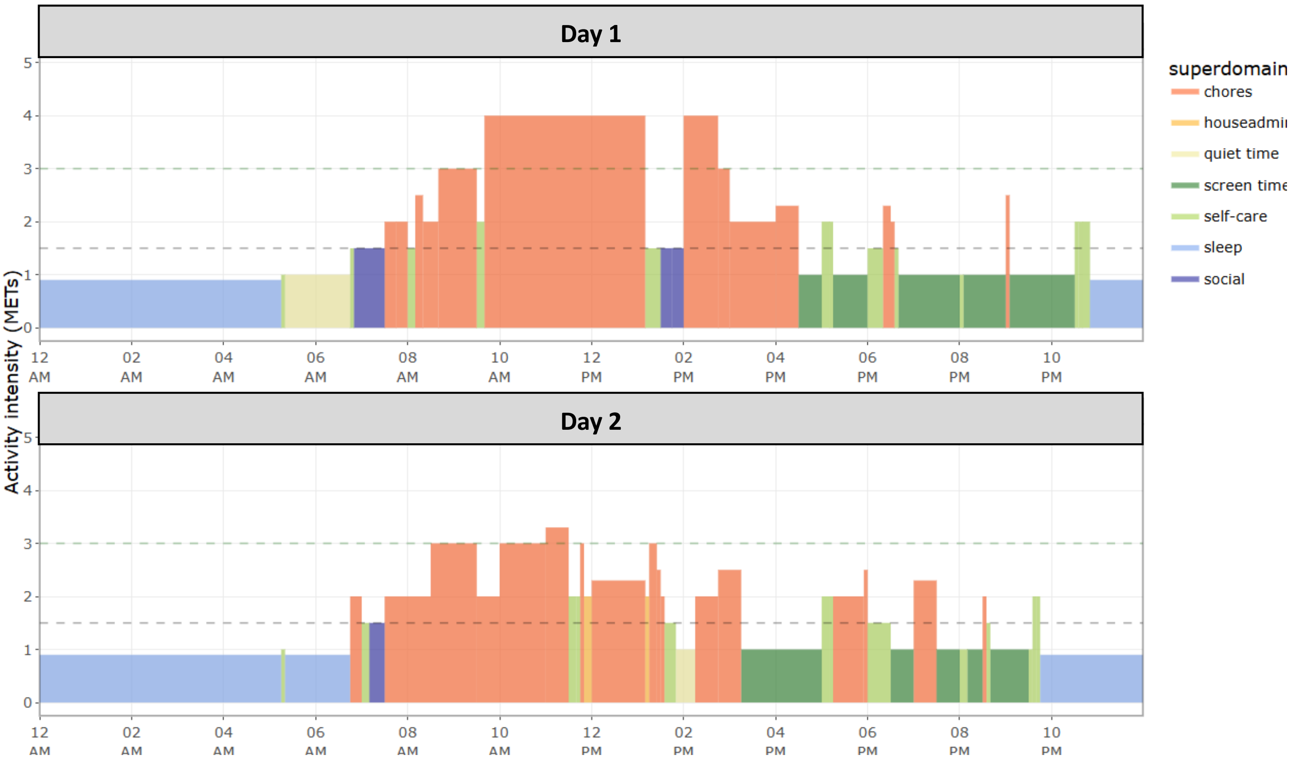

**Supplementary File 3.** The weekly check-in process

The following screenshots display the weekly check-in process that is undertaken in the Introductory phase by participants enrolled in the Extended program (i.e., intervention group). Weekly check-ins are completed independently by the participant, and are followed-up by a brief phone call with a research team member. This example displays an example check-in for a participant who has just completed Week 1 of the Introductory phase and is entering Week 2.

#### Step 1. *Reflect on past week's goal*

In Step 1, the participant is first required to rank their performance with last week's goal, from 'much less than expected' to 'much better than expected'. At the top of the screen, the participant is reminded of last week's goal (where "user example" is replaced with a specific goal, e.g., "Last week your goal was to replace taking the escalator with taking the stairs, for 5 minutes, every day. How did you go?". Participants are also asked to briefly reflect on the barriers or enablers that influenced this rating. This information is reflected on by the researcher in the phone call check-in.

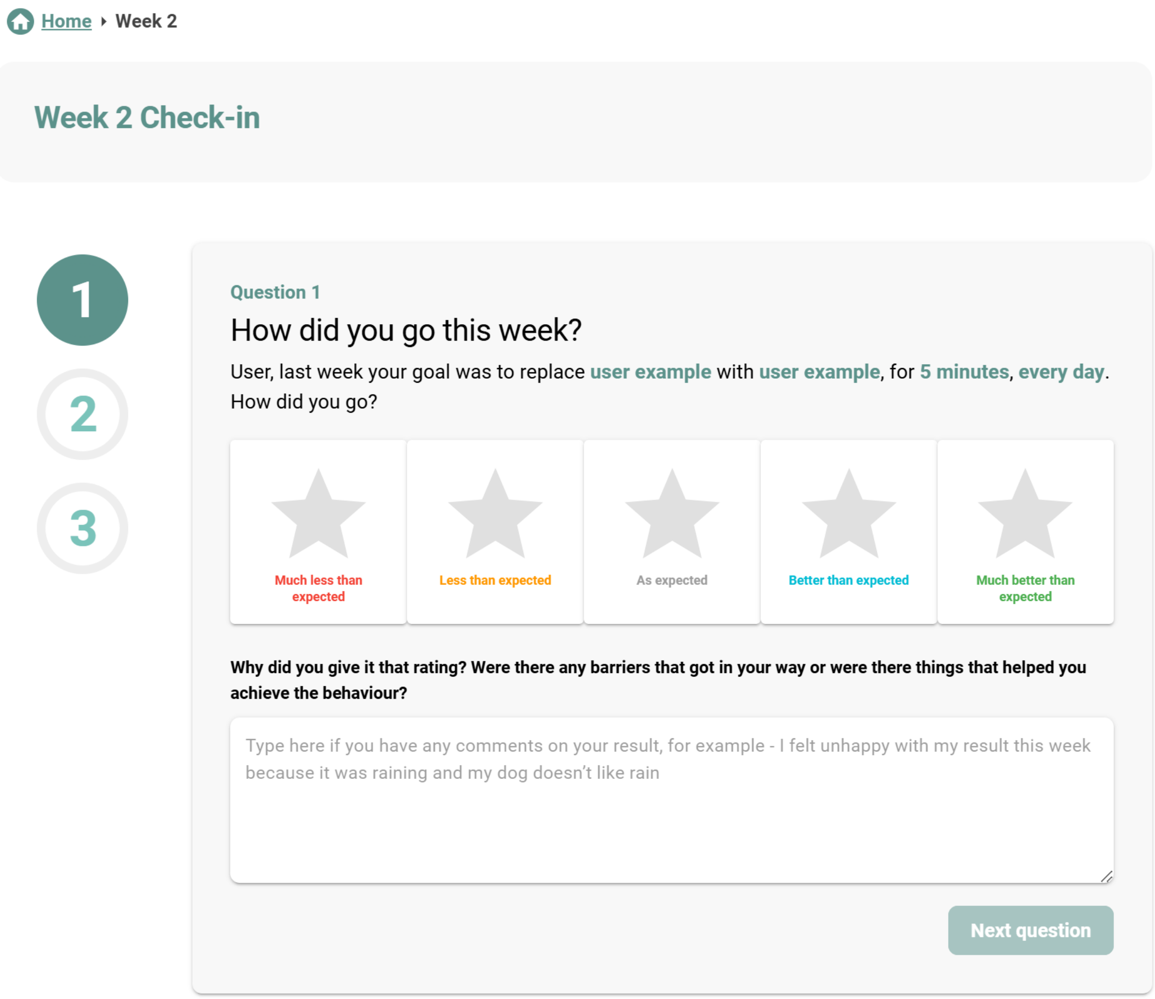

**Step 2.** ***Set the focus for the coming week.***

The participant is then asked to select their area of focus for the coming week, with choices between ‘be more physically active’, ‘improve my sleep health’, or ‘no change from last week’ (i.e., I will re-use the same goal that was set in the previous week).

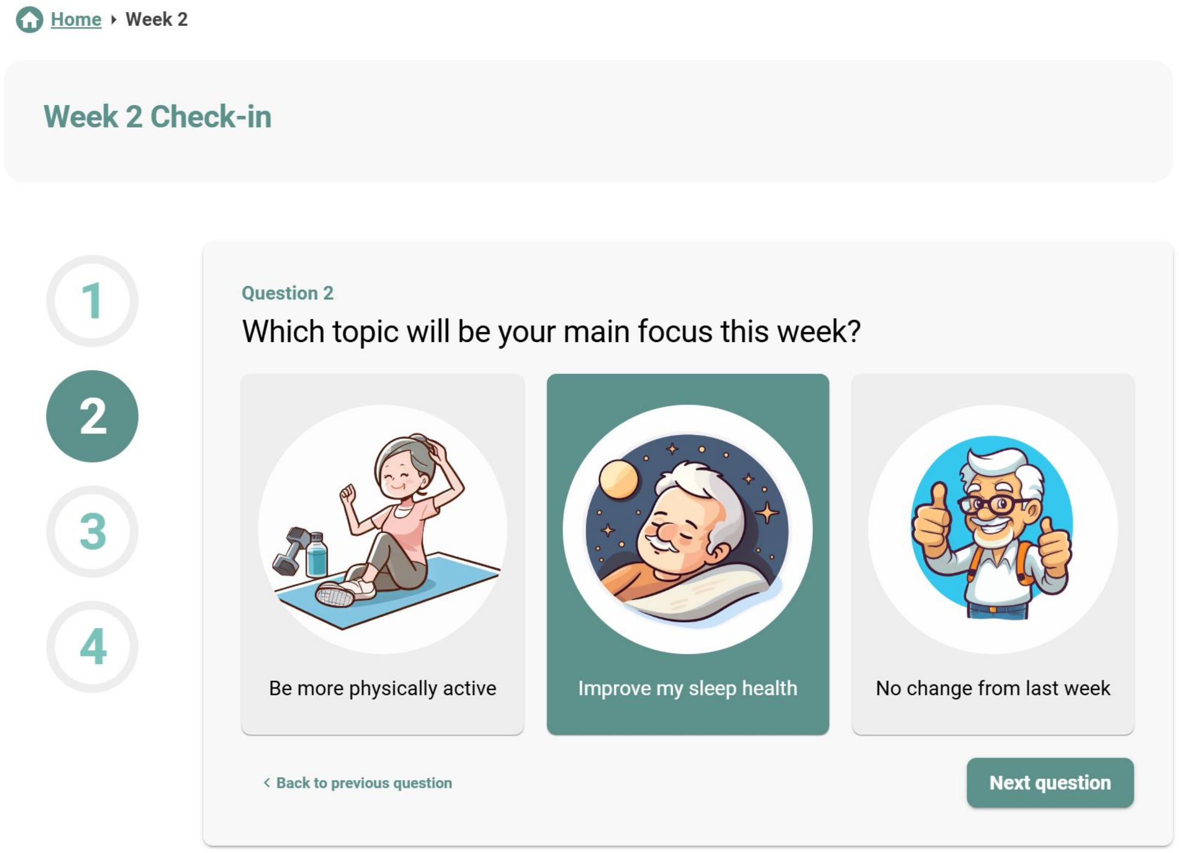

**Step 3.** ***Choose the specific goal for the upcoming week.***

Using a series of drop-down menus, participants must then select their specific goals for the coming weeks. In the displayed example, the participant has chosen that they would like to replace ‘having an irregular sleep time’ with ‘setting a strict sleep time’, every night in Week 2. Participants are asked to identify what they need to make this change, e.g., “I will set an alarm to go off at 10.00pm which is when I need to turn the television off and get ready for bed”. Finally, participants are asked to rate how confident they feel that they can achieve this goal, on a scale from 0 (not confident at all) to 10 (very confident). If participants indicate low confidence, a suggestion to alter their goal is displayed at the bottom of the screen.

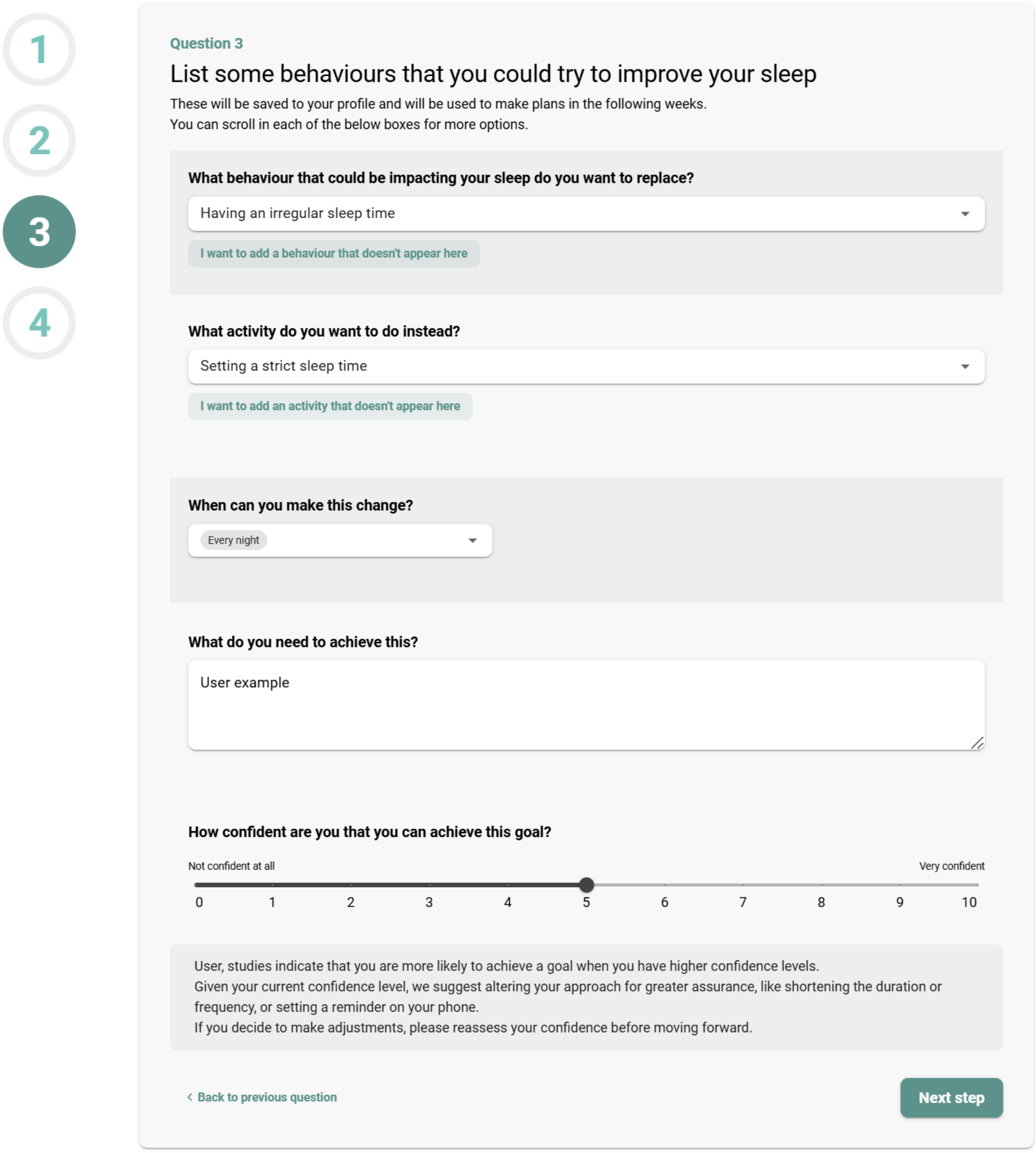

**Step 4.** ***Revise and finalise the check-in.***

At the final step, the participant is shown a summary of their reflections on last week's goal, and their selected behaviour change goals for the coming week.

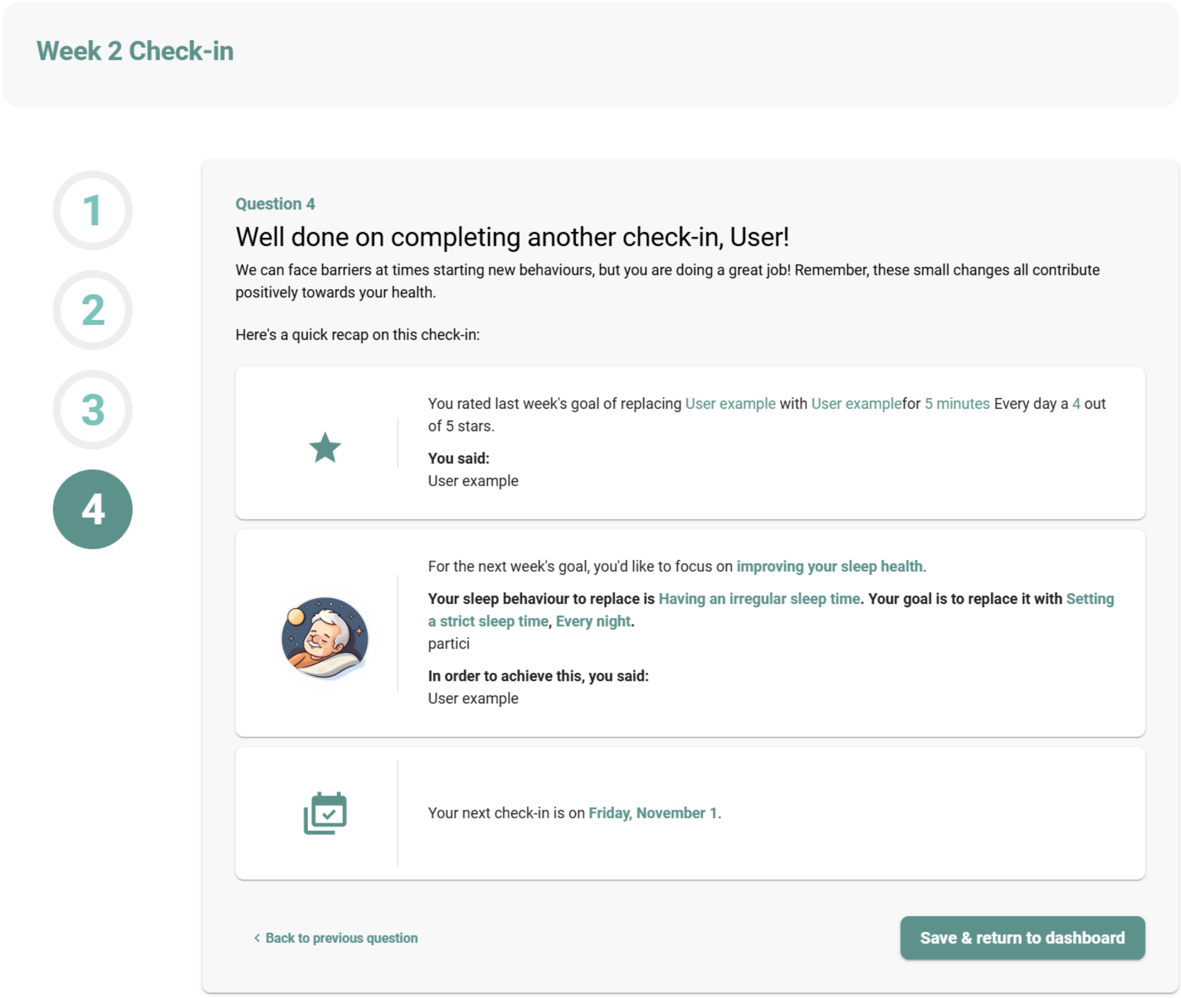

**Note:** Following the check-in, the goals for the current week are displayed on the participant's dashboard. Additionally, a targeted resource (specific to the goal of the current week, in this case, sleep) is displayed on the right hand side of the screen.

### Welcome back, User!

You are now in Week 2 of the initial 12-week Small Steps program. Keep up the great work.

Week 1 | **Week 6** | **Week 12**

A reminder of why you are completing Small Steps: **User example**.

#### Your activity plan

**Your new goal for this week**

**Replace Having an irregular sleep time with Setting a strict sleep time, Every night .**

User, remember, you stated you need the following to help you achieve this:

User example

Your next check-in is in 5 days - come back to review how you went!

**Your previous weekly goals**

Your week 1 behaviour change was to replace **User example** with **User example**, for **5 minutes** Every day.
You mentioned you could do this activity with **a friend**.

**Weekly learning**

**Sleep As We Age**

As we age, it is common to experience changes in the quality and length of our sleep. These changes may be due to mental or physical health conditions, such as depression, anxiety, heart disease, or arthritis, which can make sleeping regularly more challenging than usual. Additionally, as we age, our body's internal clock ages. This internal clock influences when we feel hungry, when our body releases certain hormones, and when we feel sleepy or alert. But did you know that in addition to being physically active there are many small strategies we can do to improve our sleep. For example, setting a regular wake time, avoiding caffeine after 2 pm, or removing screen time from the bedroom have been shown to help!

View this resource

**Your physical goal with Small Steps**

User example

**Your sleep health goal with Small Steps**

User example

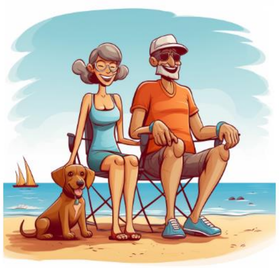

**Supplementary File 4.** Post-intervention semi-structured interview guide

| Question Category | Sub-category | Question Suggestions |
|---|---|---|
| Recruitment | Awareness | What motivated you to join the Small Steps program? |
| Health behaviour awareness | Care/guidance | Before Small Steps, where would you normally seek assistance to improve your physical activity and/or sleep behaviours? Do you feel Small Steps was different to the normal assistance you would receive? |
| Small Steps Review | Overview | What were your favourite and least favourite aspects of Small Steps? If given the opportunity, would you take part in Small Steps again? |
| | | What would you consider to be the most important aspect/s of the study? |
| | | How do you describe the amount of time expected of you to contribute to the project and the level of burden involved? |
| | Website | What did you like and dislike about the website? |
| | | If you could add any feature to the website, what would it be? |
| | | What are your opinions on the resources on the Small Steps website? |
| | Resources | Did you access the website resources?<br>Why/ Why not<br><br>How did these resources help you/support you in your journey?<br><br>Are there particular resources that you found helpful?<br><br>Or not helpful?<br><br>Do you have suggestions for how these could be improved |

| | | |
|---|---|---|
| | Phone calls (Extended program only) | Were you part of the extended program where you received weekly research support during the introductory phase?<br><br>What did you like and dislike about the phone calls? |
| | | How could the phone calls be completed to better support you? |
| | Data review:<br>Throughout the study you were provided with a review of your data... | What changes have you seen in your physical activity and sleep behaviours?<br><br>How do you feel these changes have impacted your wellbeing/day-to-day life? |
| | | We provided feedback to you about your current sleep, physical activity and sitting patterns back at the start of the program. Do you think that this information impacted your health behaviour? |
| Outcomes | Observation of outcomes | This study focussed on time use – sitting activity and sleeping, Do you feel that other health behaviours or habits could be supported in a similar manner?<br><br>If yes, What would this look like? |
| | Breadth of outcomes | Have you noticed any other behavioural, health or mood changes since taking part in the study? |
| | Scalability | If this were to be scaled up and available to the wider community, what would you feel would be the most important aspects to consider? |
| Final | Final feedback and thoughts | Do you have any final feedback for the Small Steps team that could be utilised to improve or expand the study? |
